# Understanding How the Self-Regulation in Sport Practice Survey is Received by Mental Performance Consultants for Prospective Use in Applied Practice

**DOI:** 10.64898/2026.10.29.26366811

**Authors:** Bradley W. Young, Sharleen Hoar, Lisa Bain, Joseph Baker

## Abstract

This study explored Mental Performance Consultants’ (MPCs’) perspectives on the prospective use of the Self-Regulation in Sport Practice (SRSP) survey when consulting adolescent athletes. Twelve Canadian MPCs completed mock SRSP surveys and interviews about the survey’s content and what they anticipated they could gain by using it with clients. Most of the MPCs claimed they could use the SRSP as an applied consulting resource. They believed that the SRSP could benefit the self-awareness of athletes and serve their information-gathering during case conceptualization. Most MPCs foresaw framing and enlarging SRSP statements/scores in joint discussion with a client, as part of triangulation, with some valuing the survey metrics for athletes and oneself. However, MPCs were philosophically divided on survey use, exposing uncertainty and negative preconceptions for some around quantitative assessment.

---

Mental Performance Consultants (MPCs) in applied sport psychology must gather information about a client—including their current difficulties, a profile of strengths and weaknesses, as well as their history and context, to guide an intervention. Information gathering is essential in case conceptualization [1], the process whereby a consultant formulates an intervention plan for a client. During this process, an MPC is tasked with connecting assessment to evidence-based strategies. Information can be captured through different assessment methods including client-report surveys, interviews, behavioral observations, functional analysis, performance profiles, biofeedback, and performance data [2]. The methods an MPC chooses to glean client-related information are related to their professional philosophy, their training, and whether the consulting focus is on diagnosing (evaluation) or raising awareness (coaching) [2,3].

Little published research has probed the elements of practical utility that are important to MPCs in using client-report surveys for case conceptualization. In 2001, Nideffer and Sagal lamented the disappearing literacy for quantitative survey use among the new generations of consultants, which they claimed was especially evident in needs analysis, interviewing with surveys, and delivering survey feedback to clients. Recently, Teschuk et al. [6] conducted a scoping review of four popular athlete self-report questionnaires for assessing mental skills relating to the practice setting. They concluded that there was a remarkable lack of guidance in the published literature for how to use the surveys to enhance interventions. Following a comprehensive search that returned 369 published citations pertaining to the four quantitative surveys, only nine articles detailed aspects for how practitioners/intervenors used survey scores to inform/influence ongoing approaches within an intervention (i.e., not just pre-to-post monitoring) [see 7,8,9,10,11,12,13,14,15]; only one published case study explicitly described an MPC’s use of a survey to engage a client in dialogue [16].

Altogether, there appears to be surprisingly little published literature informing how to use quantitative surveys in strategic ways within an intervention, and specifically in case conceptualization. Yet, there continue to be recommendations for more practically informed approaches to surveys and assessment [e.g., 17]. More specifically, Young et al. [18] called for fulsome consideration of how the *Self-Regulation of Sport Practice* (SRSP) [19] survey could support practitioners’ consulting approaches. They called for more applied research to understand consultants’ use of the SRSP for “the psychology of practice” [18, p.1], i.e., supporting athletes’ psychological proficiencies that facilitate skill acquisition and learning in practice settings. Despite the SRSP having more of an empirical and less of an applied origin than other surveys such as Durand-Bush et al.’s [20] OMSAT-3 and Hardy et al.’s [21] TOPS-2 [6], this call for greater examination of the SRSP through the lens of a practitioner was predicated on the notion that there is metacognitive assessment in the SRSP that differs from mental skills assessment on other surveys, and that such content might lend itself well to consultation around practice-enhancement [22]. Siekańska et al. [16] also noted there was little known about how such the SRSP survey could be used in applied practice and invited more innovation in embedding the survey in consulting dialogue. As such, this study aimed to explore MPCs’ views on the SRSP survey and its prospective use in applied practice. The current researchers chose to explore the prospective practical validity of this survey because of their scholarly interest in the psychology of practice, because some of them had previously worked on the psychometric development of the SRSP survey, and because they wished to interrogate how the SRSP might be received by applied practitioners who were apart from them.

The SRSP survey [19] is grounded in self-regulated learning (SRL) theory [23], which emphasizes the self-agency and reflexivity of learners, and outlines psychological characteristics that support how learners prioritize and direct energies towards tasks that are more likely to benefit their development. The survey features a sport practice-enhancement orientation [24]. It asks athletes to consider practice contexts in which they engage in taxing/difficult practice tasks, that are not always necessarily enjoyable, and that they consider pertinent to their talent development. Statements assess athletes’ report of various psychological processes they can enact when preparing for, engaging in, or reflecting upon their practice activities (either singular practice trials, or session as a whole) [18]. Thirty-one items assess five psychological processes related to the self-management of goal-oriented practice (see Supplementary Appendix A); items for ‘planning’, ‘checking’, and ‘evaluation-reflection’ scales are metacognitive, while scales for ‘effort’ and ‘self-efficacy for challenge’ are motivational in nature [19].

Wilson et al. [19] tendered this survey as a highly reliable and valid assessment instrument, which benefitted from about a decade of mostly psychometric endeavours in which measures that were imported from educational psychology became progressively attuned to the field of sport talent development, particularly to the sport practice setting [see 24,25,26,27]. In tracing the origins of the SRSP survey, Young et al. [18] noted this overemphasis around psychometric testing and a commensurate focus on criterion validity, which involved using survey scores to reliably distinguish athletes of varying performance levels. They further stated that the SRSP survey was “never intended to be a diagnostic test dedicated to measurement alone” [p.5] and that “one of the original intents in creating this instrument was practical validity and value to practitioners” [p.2]. Closing this research-to-practice gap therefore entails gaining active MPCs’ perspectives on whether and how they might consider using such a survey in their consultation.

Thus, our purpose was to explore MPCs’ perspectives about their potential practical use of the SRSP survey [19] as an applied consultancy resource. Considering the contemporary uncertainty for how MPCs consider surveys, we were interested in their initial attitudes towards the SRSP and the prospect of using it. The study was framed for consulting with competitive adolescent clients, an important age cohort for which the SRSP has been psychometrically tested [19] and one in which there is accelerated development of cognitive capacities and metacognitive orientations that supports intervention regarding the self-regulatory strategies of young people [28]. Three key questions were asked: i) what are MPCs’ initial perspectives on using the SRSP survey? ii) what psychological content do they think is being assessed by the survey? and iii) what information do MPCs believe they could gain by giving it to adolescent athlete clients as part of their consulting?

## Methods

### Research Paradigm

The overarching philosophical orientation guiding this study was pragmatism. Pragmatism is a practical consequence-driven view [29] that attempts to understand real world phenomena by selecting whatever combination of philosophies, methods, and procedures provides the best explanation to the research questions [30]. As such, the research aims, methods (i.e., having each MPC complete an SRSP survey for a prospective client), and recruitment criteria were all oriented towards applied findings. Pragmatism also adopts a value-oriented approach [30], which was reflected in the researchers’ and the participating MPCs’ desire to support young athletes as learners in the practice context. In essence, pragmatism oriented the study toward producing knowledge that could meaningfully inform applied practice.

Since our purpose was to understand how MPCs interpreted and made sense of prospective uses of the SRSP in their applied practice, we pragmatically adopted a constructivist epistemological lens when interpreting the interview data. Constructivism was selected because it is well suited to examining the multiple, context-dependent meanings that MPCs attribute to this topic, and for understanding how such meanings are co-constructed through interview dialogue [31]. With constructivism, knowledge is generated through interactive dialogue between participants and researchers, with the goal of interpreting how participants make sense of their experiences of a topic. Constructivists recognize that realities are socially constructed and subjective, meaning individuals actively build their understanding of the world through experiences and particularly social exchanges/processes. Consequently, knowledge and understanding of the topic are co-constructed between a participant and an interviewer.

In this paradigm, the interviewer’s knowledge/experience is not separated from the process. Rather, it can be an asset for eliciting interpretations from an interviewee, especially if the interviewer is highly cognizant of the topic, and as long as they effectively prompt the interviewee to share insights on the topic freely and extensively, and support the interviewee’s dialogue in a responsive fashion (i.e., supporting the conversation in the direction it is being led by the interviewee, while keeping it within the purview of the research aims). Notably, the first and second author are professional members of the CSPA and AASP who hold the Certified Mental Performance Consultant credential. Their professional experiences informed reflexive interpretation of the data, while their pragmatic orientation ensured that the practical utility of the findings for practitioners remained at the forefront of that interpretive process.

Although pragmatism and constructivism are philosophically distinct [29], they are compatible for the purposes of applied qualitative inquiry because both prioritize understanding experiences in context and reject the notion that knowledge can be completely detached from human experience [31]. As such, we refrained from claims of singular reality and embraced context-dependent explanations. That said, pragmatism asks researchers to consider statements on degree of agreement “as a way to approach” what might be considered practical, though not universal, truths [29, p.22]. Thus, in presenting the findings, we highlighted areas of convergence and divergence among MPCs by noting, where appropriate, the number of participants who aligned with or differed in their interpretations and preferred approaches. In sum, our pragmatic orientation justified the use of a constructivist lens because understanding participants’ context-dependent meaning-making was deemed the most useful way to address the research questions. When interpreting the results we refrained from claims of universal or objective truth (constructivism), but signalled areas of convergence and divergence in practical interpretations and approaches among MPCs (pragmatism).

### Participants

A University Ethics Board approved all procedures. All participants gave written informed consent. Using the professional roster of the Canadian Sport Psychology Association [32], MPCs were emailed invitations to complete an online questionnaire beginning November 20, 2021. Questions screened for consulting experience, frequency with which they consulted 13-18 yrs-olds, and focus on the psychology of practice. Further recruitment occurred via snowball sampling, with recruitment ending on February 21, 2022. Inclusion criteria were applied to 19 initial respondents. At minimum, participants needed to ‘agree’ to “I am interested in the psychology of how athletes can more optimally learn and manage their efforts/activities during practice” and ‘slightly agree’ to “consulting on the psychology of practice is an important part of my role.” Twelve MPCs were selected who were actively consulting adolescents who ranged in competitive level (see Table 1). The MPCs ranged in experience, and most of the experienced participants had robust adolescent clientele. Gender distribution of the participants mirrored the broader CSPA MPC roster.

**Table 1:** Demographic Characteristics of Mental Performance Consultant Participants.

| MPC | Gender | Yrs as an MPC | Clients’ Age Range | Adolescents as % of Clients | Competitive Level of Adolescent Clients | Interactions w/Adolescent Clients (times/mo) | I work with adolescents on psychology of practice<sup>*a*</sup> | I am interested in psychology of practice<sup>*a*</sup> | Psychology of practice is an important consulting role<sup>*a*</sup> |
|---|---|---|---|---|---|---|---|---|---|
| Emily | W | 11 | 10-35 | 81-90 | Provincial | 2-3 | 7 | 7 | 7 |
| Ayesha | W | 3 | 7-34 | 51-60 | Train to train, train to compete, train to win | 4-5 | 4 | 7 | 5 |
| Allan | M | 7 | 13-25 | 71-80 | Provincial | 2-3 | 5 | 7 | 7 |
| Oliver | M | 5 | 13-45 | 41-50 | High performance | 4-5 | 7 | 7 | 7 |
| Lucia | W | 18 | 12-45 | 71-80 | Elite | 2-3 | 7 | 7 | 7 |
| Anne | W | 20 | 10-35 | 51-60 | Wide range, some high performance, national, provincial | 8+ | 6 | 7 | 7 |
| Charlotte | W | 19 | 11-40 | 41-50 | Aspiring provincial & national stream, national team | 8+ | 7 | 7 | 7 |
| Maria | W | 14 | 13-60 | 61-70 | Very competitive | 2-3 | 6 | 6 | 6 |
| Myriam | W | 10 | 14-40 | 51-60 | National team | 2-3 | 5 | 6 | 7 |
| Jose | M | 22 | 10-50 | 41-50 | Regional to provincial, some national | 8+ | 7 | 7 | 7 |
| Hugo | M | 24 | 14-36 | 31-40 | Provincial/national | 2-3 | 7 | 7 | 7 |
| Ronda | W | 27 | 14-42 | 10-15 | Provincial/national | 1 | 6 | 7 | 7 |

*Note.* <sup>*a*</sup> Ratings ranged from 1 (strongly disagree) to 4 (neither agree or disagree) to 7 (strongly agree).

### Procedures

#### Mock SRSP Survey

Participants completed a mock version of the SRSP survey [19] ahead of an interview. The survey typically asks athletes about how they manage aspects of their training. Items cover metacognitive scales for *planning* (9 items), *checking* (3), and *evaluating-reflecting* (4), and motivational scales for *self-efficacy for challenge* (5) and *effort* (5). Likert responses range from 1 (strongly disagree) to 7 (strongly agree). In this study, we were not interested in gaining respondents’ self-reports. Instead, when presented with the *mock* version, each MPC was asked to “have a particular client in mind, responding as if you were said client” (see Supplementary Appendix A). They first entered demographic information for this client (see Table 2), then did the mock SRSP. This procedure familiarized the MPCs with items relevant to the assessment of psychological processes enacted in the practices of athletes. It also readied them to talk about how they might guide a client in consulting. Each MPC was emailed the mock SRSP within 48 hours of their upcoming interview and all returned it, completed, prior to meeting.

**Table 2:** Information about the Adolescent Client for Whom Participants Completed a Mock SRSP Survey.

| MPC Pseudonym | Client Age (Gender: Boy/Girl) | Client Sport | Competitive Level of Client | How long has this athlete been your client? | Frequency of Interactions with this client (times/mo) |
|---|---|---|---|---|---|
| Emily | 17 (G) | Curling | Provincial level | 3 yrs | 1-2 |
| Ayesha | 17 (G) | Soccer | Prep | 5 mos | 2 |
| Allan | 18 (B) | Baseball | University | 3 yrs | 1-2 |
| Oliver | 17 (B) | Hockey | High performance | 2 yrs | 3-4 |
| Lucia | 14 (G) | Hockey | Elite AAA | 5 yrs | 2 |
| Sofia | 15 (G) | Soccer | Major League Soccer franchise junior academy squad | 6 mos | 4 |
| Charlotte | 15 (G) | Diving 10m | International level | 2 yrs | 2 |
| Maria | 17 (G) | Tennis | US Academy | 6 mos | 1 |
| Myriam | 16 (G) | Gymnastics | International level | 1 yr | 1 |
| Jose | 16 (G) | Softball | Provincial level | 6 mos | 1-2 |
| Hugo | 17 (B) | Alpine skiing | National level | 5 mos | 2 |
| Ronda | 16 (G) | Basketball | Provincial program | 2 yrs | 3-4 |

#### Semi-Structured Interview

Each MPC engaged in a semi-structured ZOOM interview. The first part of the interview began by asking about their views on SRL, how they described it conceptually, and how they described features of effective SRL in adolescent athletes. (Responses for this segment, approximating the initial 30-35 mins of the interview have been published already [see 33] because they did not relate to the survey *per se*. These data are not treated herein.). The following part of the interview guide is the subject of the current article. The questions (see Supplementary Appendix B) asked what the MPCs thought ‘big picture’ about the survey, what psychological content they thought was being captured in it, what important information they could get from an adolescent athlete who completed the survey, what they thought the survey scores could reveal, and the importance of SRSP scores. The substance of these questions represented on average 35 mins (range 29-47) of dialogue with each MPC. The interviewer used follow-up probes to ask for elaboration or clarification on consulting scenarios.

The first author (FA) and third author independently piloted the interview guide with two MPCs. They conducted the first interview together, then debriefed to ensure coherency and to reflect on the quality of their questioning. The FA conducted seven of the remaining interviews, with the third author doing three. Interviews were recorded and transcribed.

#### Rigor

All but one interviewee were independent from the interviewers, meaning they were not personal acquaintances. The exceptional interviewee met all inclusion criteria and was a professional acquaintance. In keeping with pragmatic principles, the mock SRSP procedure served to reliably keep the survey at the fore; moreover, throughout the interviews, the MPCs were asked to keep the client affiliated with their mock SRSP in mind when answering, which focused responses on working with clients rather than abstractions. In terms of data analysis, use of the second and fourth authors as critical friends [34] promoted rigor, as they challenged the interpretation of data, its alignment with the research aims, ensuring coherent presentation of findings. The FA and second author were professional certified and actively practicing consultants, which helped to add rigor, for example, by enhancing the FA’s responsiveness during interview questioning, and enhancing credible interpretations for applied interpretations. During analyses, with pragmatism in mind, they kept an open mindset to extract novel dialogue from interviewees, which they believed would enhance the sincerity and meaningful coherence [35] of the findings for practitioners.

### Data Analysis

In keeping with reflexive thematic analysis [36], the FA began by reading all transcripts and making notes to become familiar with the data [34]. Data matching the purpose and our key questions in this article amounted to 68 pages of single-spaced transcribed text, which were coded apart from data for Young et al. [33]. The FA initially coded passages at a lower-order, semantic level, populating lower-order themes into separate coding sheets corresponding to the sequence of the interview. Within each sheet, the FA inductively analyzed the data to create middle-order themes of shared meaning. The FA next met the second author to present the sheets. They elected to collapse sheets from the early interview segments (i.e., ‘big picture thoughts’, ‘global sense of what is being captured’, and ‘what psychological content is being assessed’) into a higher-order theme for ‘initial reception of the survey and its content’. Second, they noted a latent, cross-cutting theme involving tensions between different MPCs around the use of a quantitative survey, which warranted its own higher-order theme. Third, we made the latent observation of distinctions in the data for the information that MPCs could acquire by giving the SRSP survey to a client. Specifically, substantial data represented what an MPC could learn using the SRSP to initially gather information when figuring out a client’s case, whereas other data represented how an MPC believed they could begin gainfully acting on the SRSP information in joint work with a client. Each of these interpretations warranted their own higher-order theme. After interpreting the higher-order themes, the FA sequenced them in a story-telling fashion, beginning with how the MPCs initially received the survey and its content, then ordering them according to phases of case conceptualization (i.e., an MPC initially gathers information to understand a client’s case before acting on this information), while limiting duplicity among subthemes (see Supplementary Appendix C for a table of themes and subthemes.) All MPCs have been given pseudonyms (Table 1).

## Results

Findings begin with the MPCs’ perspectives upon initial receipt of the SRSP. We then present philosophical divisions among the MPCs around using the quantitative survey, which emerged early and influenced their subsequent responses about the SRSP. Next, findings are presented for what the MPCs anticipated they might gain by giving out the SRSP: first, for their own benefit in an information gathering role; second, for acting on this information by extending from the SRSP to develop proficiencies in clients.

### The MPCs’ Initial Reception of the Survey and Its Content

The MPCs first shared ‘big picture’ perceptions on the survey and their perspectives on the type of content being captured. All of the MPCs, except one, were naïve to the survey prior to being emailed. They disclosed several notable subthemes, including: the SRSP can elicit information benefiting the MPC; the survey can elicit greater self-awareness and help structure self-feedback for athletes; its content captures the extent to which an adolescent gives themselves a role in their own learning; it captures SRL, metacognitive and motivational content; it neglects to assess some content.

Many of the MPCs first remarked that the SRSP could elicit valuable information. Lucia saw it as useful “to get to the nitty gritty in what [athletes] are thinking and feeling during practice—this survey kind of helps me to see whether they’re actually doing that [psychological processing]”. She believed that information generated from the SRSP would allow her to move past unreliable assumptions she might have about what athletes understand in terms of cognitive skills around practice. In the absence of a similarly pertinent survey, Maria believed the SRSP could meet an applied need, commenting, “This is one of the places where we have to grow as MPCs. We need to be able to generate some sort of evaluation or assessment of growth around learning and practice.”

The MPCs initially responded that the survey could raise self-awareness benefiting athletes. Allan explained that when an athlete responds to the SRSP, “they’re getting an understanding of how well they process information in their practice and ultimately providing themselves feedback with how well they’re doing.” He said, “part of this is having an athlete understand the perspectives they can have in their own evaluation.” For Sofia, the SRSP “actually asks an athlete, ‘are you thinking about what you’re doing?’ and invites them to engage in “a process focus” when appraising their practice habits. Ayesha contributed, “having a client do the SRSP would be a great self-awareness exercise around practice. The questions challenge them to think about themselves, like an entire reflective process.” She explained that a process supporting self-awareness and self-feedback could be approached in a more structured fashion using the survey. Ayesha reflected, “the SRSP would be perfect for working with a client who’s like, ‘I’m not getting better, and I don’t know what’s going on’. So, really creating some sort of structure for them to approach practice in a way that allows them to be mindful and deliberate.”

Several MPCs noted that a more structured approach to enhancing awareness on psychological functioning around practice would meet a need, because this topic is not prioritized for adolescents. For example, Maria explained:

> Coaches don’t spend any time explaining the things exemplified in this survey to kids about how they can manage the practice situation more effectively, how they can prepare for it, how they can manage their emotions, the distractions…That’s what gives the SRSP some real value.

Allan opined, “young athletes obviously understand, like, how a practice is important. But they won’t apply as many mental skills in practice, or as frequently, as they do in competition.” The MPCs appreciated that information from the SRSP would signify an important focus and provide new insights on practice. Whereas Sofia thought such focused self-awareness could benefit adolescents mostly in preparing ahead of practice, many of the MPCs saw “more self-awareness and self-regulation around these pieces [in the SRSP] as an opportunity to increase the quality of practice” (Allan).

Some of the MPCs remarked that the survey content would capture the extent to which an athlete gives themself a role in their learning process. For Oliver, the SRSP:

> … surveys an individual’s ability to take learning in their own hands and understand themselves and their performance enough to be able to make adjustments wherever necessary. [The SRSP] is capturing, quite well, self-regulated learning as a concept, and as a cycle of reflecting, preparing, and performing…But it pushes a little bit further. There are certain concepts of motivation, and some autonomy surveyed for sure, and being open to applying yourself to that learning, and the athlete actually having a role in that. Which, unfortunately, athletes don’t really have a say all that often. So being able to foster that is important.

Use of the survey could support adolescents’ development of competencies whereby they can more confidently insert themselves into their own learning. For example, Ronda said, “athletes [knowing their responses on the survey] could really create some self-awareness that they too have the capacity to engage in their own learning”, which feeds the notion that “they can plan their effort, they can control how much effort they put in, they can evaluate themselves. Knowing that I can evaluate myself, that I don’t have to always rely on feedback from my coaches, that’s a big [insight]”.

The MPCs commonly highlighted three categories of content reflected in the SRSP. All commented that “the SRSP invites athletes to look at like how well they apply self-regulated learning processes in completing tasks or [working on] technique.” They all recognized that the SRSP comprehensively captures metacognitive content important to practice and many acknowledged that the survey effectively represented motivational content. A few of the MPCs discerned what the SRSP did not capture. Two MPCs felt emotions were not portrayed directly and could only be addressed if they were to impress their own narrative on emotional responsiveness onto the items. For example, Charlotte pointed to “[item #12] ‘something unexpected’ or a coach changing the planning [on the athlete]”, adding, “this item definitely relates to the reality of training. And so how does the athlete react to that? And there’s going to be emotional responses first that need to be considered, before I consider the athlete’s ability to adapt and re-plan.” Ronda commented, “The SRSP is reporting how much the athletes use self-regulated learning strategies or do those strategies. If people are using these strategies more, they get a higher score, though it might *not* capture if they can do the strategies”.

### Divisions Among MPCs Around Using a Quantitative Survey in Consulting

How the MPCs viewed what they could learn by giving out the SRSP depended on their philosophy. Several MPCs preferred to figure out and set the guiding questions in client cases based predominantly (if not exclusively) on interviews or narrative approaches; these MPCs were more likely to problematize the survey or mention limitations to its use early in the interview. Notably, their issues with survey use were not exclusive to the SRSP but extended to any survey. Their responses portrayed tension between survey use for diagnostic assessment and narrative intake.

For MPCs who philosophically preferred narrative approaches, their responses indicated a hesitancy to embrace surveys. This was illustrated by Myriam: “I’m not sure I need to administer the survey to know gaps, what needs to be worked on with a client. I prefer interview techniques and just ask the same questions.” She added, “I can build rapport more in the conversation. Particularly with adolescents, there’s so much variability, I avoid any type of survey. And I’m not sure how accurate a survey is.” Allan cautioned, “You can go overboard with surveys. So, I guess I would have more questions about how this all fits in the bigger picture, in terms of like assessment within my [consulting] practice. Like, I kind of limit assessments.” Jose noted a “personal bias when it comes to assessment”, adding “I let athletes look at surveys and don’t even score them. Like, let’s just talk about this questionnaire, why it matters and how you can integrate these kinds of thinkings into your preparation.” He worried about “using survey assessments to pigeonhole people into boxes with scores.” Emily said, “I wouldn’t use any survey with some athletes I work with.” She offered, “I’d ask some of those questions [from the SRSP], but face-to-face so I could get more information and ask follow-ups. Like, ‘are you feeling like you’re reaching…?’, ‘are you able to adjust…?’, ‘were you able to achieve…?’, like, ‘how are you feeling?’.”

Several MPCs were concerned that surveys could be construed as a way of “testing” a client and as a form of “assessment” for diagnosing weaknesses. Myriam, Jose, Allan, Emily and Ayesha, who were in this camp, cautiously viewed any survey as an impersonal diagnostic instrument. They revealed they rarely used surveys; they shared philosophical reasons for alternative methods, and worried, for example (Ayesha), about “taking a survey and using it as an independent source to define a person”. Ironically, when these MPCs were asked later in the interview whether the SRSP scales addressed what they were supposed to address, their responses fixated on validity and reliability of scores (i.e., psychometric terms) despite professing their disinterest in psychometrics. Instead of talking from a narrative perspective, they talked about “accuracy” and how they were uninterested in “reliability and validity” of instruments, in some cases sharing doubt about the accuracy/skewness of self-report scores. Thus, they fixated on a view of surveys as diagnostic tests, disjointed from humanistic approaches to understanding a client, with a risk of scores casting respondents in a critical/unconstructive light.

Other MPCs were in a different camp. Maria, Hugo and Ronda were devoted to surveys and saw them in a constructive light. They often used surveys in strategic, reasoned ways to enhance their client interactions. These MPCs were quick to say that they find surveys helpful in furthering dialogue and find that adolescents like surveys and putting numbers to things. Curiously, in later segments of the interview, when they were asked whether the SRSP scales and items addressed what they were supposed to, their responses focused on what they thought of the content of the items. They were less likely to stray into conversations on score accuracy, validity and reliability. A couple of these MPCs acknowledged that they knew about all the work that goes into establishing the psychometrics of a survey but added that this was not really a concern for them when using surveys in applied practice. The three other MPCs—Lucia, Oliver, and Charlotte, did not appear to fit in either camp and liked to move between survey use and narrative case examination, depending on context, client, preferences and needs.

Altogether, there was an ironic tension impressed on many responses for how the MPCs would receive the SRSP, depending on the camp in which they presented themselves. Those who embraced surveys did so because they intertwined assessment with dialogue. Those who were dismissive or highly reserved saw survey assessment as impersonal and curtailing dialogue. As these MPCs worked through the interview segments dedicated to the SRSP survey, this tension somewhat diminished, with all the MPCs eventually acknowledging ways they could see prospective reasons for using aspects of the SRSP.

### Using the SRSP to Initially Gather Information to Understand the Client’s Case

The MPCs portrayed different ways of using the SRSP to initially gather information about a client. A prevalent subtheme was that the scores mattered as a source of information they could analyze and interpret to understand a case. Although they varied in their convictions about the absolute importance of numeric scores, most of the MPCs saw lower scores (or unusually lower ones) as flagging an area to interrogate to determine whether there was a need for improvement. They expected to spend more time in dialogue in these areas. Maria said, “If I see a 4 when it’s typically a 6, we’d have a conversation about that.” She would “be honest that we’re low, ask them why they felt that way” because this score tells me that something’s happening, they’re not leaning in, not adapting, or not managing the challenges.” Ronda added, “I’d find out what sort of frustrations they’re facing [around a low score], what [practice] tasks they didn’t like and what they didn’t like about them.” Most of the MPCs wanted to figure out “whether one scale is a bigger issue than another” (Lucia). Rather than levels of scores, Emily would inspect whether certain SRSP scales show more variability and “try to understand what those fluctuations are about.”

Myriam had a distinct perspective on gathering information by exploring low and high scores. She conjectured that attending to lower scores might tell her that a client lacks knowledge/support for developing SRL skills. She explained, “I need to figure out if they’re low because they’ve never been exposed to this content, like, maybe they don’t know that this is gonna help them”, adding, “maybe their coach never gets them to do anything like this, like, they just go to practice, do as their told, and don’t think about it.” Alternatively, she anticipated that she might be able to verify whether high scores represented initiative by an athlete: “I would try to figure out, is this something you work on with your coach, or something you’ve done yourself? And how did you come to do that on your own?”. In doing this, she felt that she could determine whether high scores might be a consequence of obligation because a parent, coach, or MPC had asked the athlete to engage in SRL content.

Overall, the MPCs said that the scores would matter because they could help determine areas for improvement and athlete learning. There were several other subthemes for how the MPCs proposed using the SRSP to analyze and interpret information for their own understanding, including: to determine the extent to which an athlete is cognitively engaging in their practice; to determine a client’s intentionality and coping tendencies; to gauge aspects of ownership, commitment and self-motivation; and, to better understand constraints on self-directedness within an athlete’s training environment.

Several MPCs believed that the SRSP could be used to tell them whether an adolescent athlete is taking the time to really think about their practice activity. Lucia said, “Because most of the SRSP items are related to goals and actions, and whether the athlete reflected on them, responses will give me some key indication as to whether they’re really thinking about their activity, that they’re not just going through the motions.” She added, “It will give me an idea whether they are actually learning to self-regulate, which is invaluable; we want them to be able to do that, especially at this age, to develop that autonomy.” Sofia said the SRSP could tell her whether a client “has a pre-practice preparation” and “a post-practice evaluation”, or “whether they’re [more laissez-faire], you know, like it’s only in practice, and then they leave [practice], not reflecting on it at all”. Lucia contributed, “if the athlete is not really taking time to think about their practice, then the SRSP helps us as MPCs as one of the tools that we need to give them when we are searching for ways to help them develop those skills, and to apply them.”

Two MPCs described how they could work through the SRSP with a client to glean insights into an athlete’s intentionality and coping. For example, Hugo said, “first, I could use the survey [to get a sense of] the intentional aspect of the athlete’s approaches to practice. Like, what is the athlete planning to do going into a session, what is the training session about, and how do they prepare for that”. He continued, “it would also be interesting to get into the coping part. Like, when the athlete hits challenge, when it becomes difficult, how do they deal with that?”. He explained his interest in using the SRSP to move into this area because “it’s natural that athletes bring up a general question first, like, ‘how do I deal with this, how can I meet this challenge, in terms of ‘how can I cope?’. I see that reflected in the SRSP.” Emily thought that she could use the SRSP to get “an idea of the athlete’s perception of their ability to learn from non-achievements of goal pursuits during practices, and their perception of being able to cope and adapt along the way.”

Some MPCs perceived that they could work through the survey to gather information about an athlete’s ownership, commitment to learning, or self-motivation. “Because it is first-person oriented”, the SRSP would allow Hugo to gauge “ownership by the athlete in terms of self-regulated learning processes.” He described how younger athletes do not yet know how to apply these skillsets, that “they’re trying to figure this out as they go. And so, supporting them with that, whether it’s an assessment or a survey, or whether it’s some consulting, will help increase their feelings of independence, their ownership”. Oliver shared that the SRSP might tell him about “an athlete’s commitment to learning, growth and development” and an adolescent’s “willingness to play a role in learning about themself and learning new skills and applying those new skills to getting better. And their willingness to see how maybe we can apply those skills in a more effective or efficient way.” Myriam said that the scores on an athlete’s SRSP “would show me how driven they are”. She pointed to items #26 [‘clearly plan my course of action before starting practice tasks’] and #27 [‘look back to see if I did the correct procedure’], offering, “anybody who’s doing those things is highly motivated to improve.”

Finally, Myriam proposed interpreting information from the survey to determine whether the athlete’s context allows for independent, autonomous activity, i.e., self-directedness in training. She explained, “So, the SRSP questions would be conducive to identifying, is it an independent type of environment or not for the athlete? Because, you do sometimes see in larger group training where they have to be more independent, because they’re not getting one-on-one attention [from the coach] that they might have in smaller groups.” She claimed she could use the SRSP to figure out the extent to which an athlete was directed by coaches relative to the opportunities for that athlete to insert themselves in managing/initiating facets of their own learning.

### How the MPCs Could Begin Acting Upon the SRSP Survey to Formulate an Intervention

In addition to describing the useful types of information that the SRSP survey might permit them to gather about an athlete, the MPCs also relayed how they would begin to use the SRSP to formulate an intervention. Specifically, they discussed psychoeducation and moving the dialogue towards the development of psychological proficiencies according to four subthemes: i) starting a discussion by framing SRSP statements/scores for a client; ii) using the survey as a resource at intake, then corroborating; iii) enlarging the survey items in joint discussion; and iii) using survey scores as metrics for the client and for oneself.

#### Starting a Discussion by Framing SRSP Statements or Scores for a Client

All of the MPCs explained that it would be critical to tie the SRSP to discussion. For the initial skeptics of surveys, like Jose, “the SRSP is foremost a discussion tool”—they demanded this to justify using the survey. Emily said, “I can put the SRSP in my consulting toolbox because it will help guide questions that I use in my narrative approach.” It was necessary for an MPC to frame survey responses thoroughly in a joint exchange where they could supply more information to a client as needed. Without this tactic, Emily explained that most adolescents would look at their SRSP responses and not know what to do with them. Myriam noted that surveys can be tricky with adolescents because of short attention spans, poor comprehension, and teens feeling they are being tested. By framing the SRSP survey content within a discussion, Myriam could build rapport and confidence in connecting her interview/narrative intake approach to subsequent facets of intervention.

Charlotte commented that MPCs generally need more methods to lead young athletes to find their voice during consulting sessions. She explained:

> The individual is most important, not the numbers on that survey. In observing how they’re responding, can we create spaces [as MPCs] where the athlete answering the SRSP question asks questions? Can we create a space where they feel comfortable in elaborating and asking, ‘well, I think this is fine…but…’. Like, creating a dialogue that opens that up?

The MPCs felt that they could frame discussion to engage the athlete’s voice by leading the client to ‘question the questions’. Jose said, “I’m less interested in actual scores and more interested in discussing afterwards why these questions are asked and what they mean for the athlete.” He continued, “The SRSP is a really good educational tool to talk about together, to work through the items, like, ok, ‘We see how you scored in these areas, but why do you think those [survey] questions are important to your training?’.” To ensure rich back-and-forth dialogue, Myriam talked about using the survey as a ‘think aloud’ tool. While sitting beside her 16-yr-old gymnast as she completes each statement, Myriam explained that “we’d still have to pick [Likert] numbers as we go through the survey. But that wouldn’t necessarily be assessment. It wouldn’t be a summative thing, we’re not adding these scores up. It would be more so, ‘we’re going to identify gaps together’.” She was keen to look at the SRSP items to support a discussion wherein the client recognizes strengths and weaknesses (gaps).

#### Using the Survey as a Resource at Intake, Then Corroborating

The MPCs discussed potentially using the survey in relation to client intake and taking steps to corroborate information from the SRSP with other sources of information/assessment. Hugo liked the idea of having an athlete’s SRSP responses ahead of intake. Before meeting with a client, he anticipated he could scan the survey results and plan out an interview schedule. He said he would “start by writing down my own questions, ok, for each SRSP scale, this is what I would want to figure out or know about a client. This is a question I can ask.” He wanted to “create a semi-structured interview, post-survey, so I can go through the results and make sense of the athlete’s scores with them.”

Jose cautiously regarded use of the SRSP around the time of intake: “I don’t usually use surveys in a first meeting, probably not even second or third one.” He prefers to first really get to know the client “by asking a lot about their sport, ambitions, history, successes, failures…and even bigger picture things”, refraining from “jumping right into assessments or surveys”. However, he explained that should a client present concerns related to “ineffectiveness around practice, that they are not making progress, or they’re getting frustrated in practice easily”, he would “bring the SRSP in…to discuss what matters for practice habits, what they are or aren’t doing around that.”

Maria saw the SRSP as a valuable “a jumping off point, perfect for my intake” and even suggested she would have a client complete it ahead of the first session:

> It would give me some info about what I should focus on. And I’m very confident in my own ability to pick that up [and corroborate that info] in a conversation. But I would validate the information we get for what to focus on from the SRSP from several sources, so, triangulation.

At another point, however, Maria explained that she would not select any survey without first determining the need for it for a client. Thus, there are times when she might meet with clients before administering the SRSP, meaning that survey-related strategies would corroborate other, earlier observations.

Several MPCs noted that the survey is but one source of information they could act upon. Emily stated, “The SRSP is valuable as one source of info, and with follow up. Ayesha said, “There’s gaps in questions on assessment [tools]. I’m not confident with this survey alone to get a good visual for my athlete.” Allan said, “self-report data are useful to compare to some other data you get. The survey is one perspective, it’s all data that’s relevant, to get an understanding of skills and how athletes tend to react to different situations.” Thus, the MPCs noted that the nature of a joint conversation with an athlete could be based in part on the SRSP, though information would need to be triangulated in other ways, including observations and coaches’ feedback.

#### Enlarging the Survey Items in Joint Discussion

Several MPCs expressed that they could take the SRSP statements and enlarge them in subsequent exploration with a client. Specifically, they perceived that the SRSP items would permit the broadening of joint dialogue about the person, their context, and important training scenarios. Enlarging the survey items was associated with two subthemes, including the goals of facilitating a client’s self-awareness: a) of difficult practice scenarios, and b) of what matters, then motivating them to work on those matters. With respect to enlarging, Myriam stated:

> I don’t need to add any items to the SRSP, I’d use them as they are, to dig deeper in a narrative. Like, if the items get too specific, then they’re not general enough for me [laughs], they’re not allowing for the nuance of context and culture, the nuance of the environment and whatnot…If they get too specific, they get controlling and are no longer general enough to be able to encompass a lot in conversation. And my conversation is going to be a lot richer than a number on a scale.

Myriam and Ronda complained that statements in other surveys are too exact/precise and thus they constrain opportunities to enlarge dialogue. Hugo commented that he thought the SRSP items are “just right”, allowing him to exercise his “prerogative” to use and “step through” the SRSP items to lead discussion in his own way. With respect to the person, the MPCs anticipated that they could enlarge discussion beyond the scores to consider the athlete’s perceptions of confidence, self-leadership, competence, personal preferences in using SRSP processes (e.g., planning, evaluating-reflecting), and an athlete’s perceived capacity to improve these processes. To do this, Ayesha said she would need a visual summary of a client’s responses, broken down for each of the five scales, that she could access before consulting. Charlotte suggested developing a bank of questions related to each scale that could be used to pull out richer information.

#### Facilitating a Client’s Self-Awareness of Difficult Practice Scenarios

Several MPCs foresaw acting on the SRSP content to help an athlete develop self-awareness about scenarios of difficulty. At the conversation-starter stage, in the ‘think aloud’ procedure with her gymnast, Myriam would try to “get at context”. She wished to question the gymnast about different scenarios that were difficult during training and even suggested adding a section to the survey where the client could identify these situations.

Allan stated, “The SRSP just provides good context for discussion points about things that don’t work for them, things that they bring up, or mental performance issues they’re having around skill acquisition.” To help the client make more meaning of their practice difficulties, Hugo said it was “his job to tailor the SRSP and the conversation a little bit towards the kind of mental demands that are placed upon the performer in a particular sport and the use of self-regulated learning skills that come with that.” He explained that knowing how his 17-yr-old alpine skier’s training demands differed from a curling client’s demands was integral for determining how he could leverage the SRSP. Allan believed that effective joint exploration that enhances an athlete’s self-awareness of difficult practice scenarios could lead to “insight into other kinds of potential barriers to practice performance” and steps to develop skills for managing confidence, emotions, and inflexibility when things are not going well. He also believed that enhancing athlete self-awareness via a scenario-focused process could lead to other benefits, such as the client coming to understand “the context for how I can support them as an MPC.”

#### Facilitating a Client’s Self-Awareness of What Matters, Then Mobilizing Their Intentions to Work on Those Matters

Three MPCs saw the SRSP as a resource that could be enlarged in a two-step process—first, by raising a client’s self-awareness of what matters to work on, then incentivizing psychological skill development on those matters. Ayesha, for example, said any consulting exercise would involve “the athlete and I discussing early on what may be valuable to work on. Like, we can recognize from the SRSP if the athlete lacks awareness that they don’t have a pre-practice planning approach.” She would facilitate dialogue to help the client see this gap, adding, “then we could discuss whether we think [filling] that [gap] would be valuable. And it may, or it may not be, right? That’s why it’s important to connect that awareness and value.” Hugo wanted to attend “to lower ‘hanging fruit’, or lower scores, to go into conversation, because those are areas where we can start to make a difference, where the SRSP lets an athlete know where they can make improvements.” He would ask a client, “ok, this is what the SRSP tells you, but do you recognize that?” or “is there anything that jumps out, or you’re surprised about [from your scores]?”. He said, “that’s not taking the completed survey as truth, it’s prompting reflection [by a client] on how they completed the survey.”

For Allan, shaping the conversation towards what matters would mean asking the client about “how the scores they gave on the survey fit their experience.” Allan would encourage the athlete to elaborate, by querying: “Well, these scores can show us how you react to different situations related to self-regulation in practice—what do you think about that? Do you have examples of that?”. Myriam foresaw using the SRSP as part of a “foundational, self-awareness thing I always start with all athletes.” Yet, instead of posing questions to raise a client’s self-awareness about fundamental mental skills (e.g., “Do you use goal setting for practice?”), she proposed asking about an SRSP process, such as planning. By asking, “Do you think about planning? Why or why not? If not, have you ever thought that it could be helpful?”, she felt she could help the client discern whether this process mattered and incentivize working on it.

Proposing his own two-step strategy, Jose would first shape dialogue with his 16-yr-old softball player so that she vocalizes where her lower numerical values are on the SRSP. He added, “once she knows these values, the next step is to have her challenge these scores.” He would initiate a conversation where “habits that have gotten her to the provincial level will be challenged, to prepare her as she transitions to the National team”. Ronda anticipated initially inviting her 16-yr-old basketball client to appraise her survey responses and getting the client to rank herself using the scores from the different SRSP scales. Ranking the scales for which she is stronger or weaker is “a way for her to know how to specifically better engage in her learning.” Next, Ronda would ask, “How do you think an elite player in your sport would score on this [e.g., checking]? What do you think they would be saying?”. She would explore the “difference from that to where you are now”, prompting, “What do you need to do to get there? What would that look like?”. Ronda believed this approach could bring awareness to gaps, while permitting the client to bring in different/new training behaviors and encouraging them to intentionally work on psychological proficiencies. Ronda also felt that this approach to having a client project their SRSP profile to a more elite level could encourage discussion on growth mindset: “I’m trying to use the scores to get the client to realize, ‘oh, other really elite people get there because they train hard, not just because they’re gifted’.”

Altogether, the MPCs presented several ways to potentially enlarge the survey items in an early phase of joint dialogue with a client. They believed enlargement served subsequent steps in their intervention, framing “areas where we I can plan my intervention” (Charlotte) including subsequent strategies to support psychological skills training of specific methods or modalities (e.g., goal setting, visualization, biofeedback training).

#### Using Scores from The SRSP as Metrics for the Athlete and for Oneself as an MPC

Four MPCs disclosed that they really liked using survey scores in strategic ways to advance their interventions. These MPCs could see the value of the SRSP scores for giving athletes a metric for self-monitoring over the course of a consulting trajectory. Maria noted that adolescents enjoy putting numbers to things: “I have a rating practice where we read everything [together], we rate practice in terms of confidence, in terms of knowing. They understand the first thing I’m going to ask them is a number on a scale.” Maria explained that, in addition to using these numbers to corroborate what the client says during consulting sessions, she would encourage her adolescents to score themselves at different points going forward. Hugo would use the scores early to set “a baseline.” Sofia voiced that scores “give athletes a valuable barometer” that they could use to track themselves at repeated points across time, which allows athlete self-appraisal of development on specific psychological proficiencies.

These same MPCs who liked numbers also described how they could use an athlete’s repeated SRSP scores to monitor the direction of their intervention and to gather evidence for whether the client is improving on certain proficiencies. Charlotte said, “The SRSP scores would let me see the impact of my work, to monitor if I made a difference.” For Lucia, the scores could help her “get to the bottom of whether these skills are being learned and applied by the athlete.” Emily and Ronda noted that trending toward higher scores over time could provide evidence to validate their work as MPCs. Picking out improvements on scores could be important for validating an intervention, however, Ronda made a final comment so as not to overstate this point---“qualitative learning happens even when an athlete gives the same score over time”, which is something to which MPCs should be attuned.

## Discussion

Our overriding purpose was to explore MPCs’ initial perspectives about prospective use of the SRSP survey among competitive adolescent athletes. In our sample of Canadian MPCs who work on the topic of psychology of practice, most of the responses were favorable. Most interviewees were open to considering the survey as a resource or, at least, adopting facets of the survey that they could tailor to their personal approaches. There was, however, an initial tension where five MPCs were reticent to integrate the survey into quantitative assessment practices, which appeared to stem from broader perspectives on survey administration, and which initially constrained what they thought they could gain from the SRSP.

### Views on Assessment with Quantitative Surveys in Applied Practice

The term ‘assessment’ pertaining to quantitative surveys appeared to be attached to negative preconceptions for these five MPCs. For this group, assessment could be narrative and dialogue-based, where they wished to rely on their own generation of questions and personal checklists of queries to engage a client in qualitative description during case conceptualization. For them, assessment with quantitative surveys was associated with testing, with clinical insinuations involving diagnostics that seemed laborious, or with a psychometric world that was somewhat foreign or not applicable to them. The impertinence of quantitative surveys, including the SRSP, appeared related to their professional consulting philosophy, which is a critical factor guiding practitioners’ selection of assessment methods [37].

This camp of MPCs never did explicitly claim that they would use the SRSP. However, we observed that they did, throughout the course of the interview, describe conditions under which this survey could be used. They articulated circumstances in which the SRSP could be employed, for example, when they had already used dialogue to ascertain that there are trying situations compromising an athlete’s ability to train independently. They also described instances where the SRSP could complement their narratives, for example, by lifting specific statements from the survey (not the whole survey or not even entire scales) and plugging them into ongoing dialogue with a client. These applications/derivatives of use contrasted with their initial comments, which seemed oriented broadly against quantitative surveys. Moreover, using surveys as derivatives—in essence, ‘plugging and playing’ items into a consulting narrative represented a personalized and non-traditional way of using quantitative surveys.

Considering there were other MPCs who wished to use quantitative surveys as they were designed, as multifactorial assessment tools, one of our notable interpretations is that assessment is not a monolithic practice or term. Our results showed that it can be complex, situated within conversational narratives, leaning on interpretive extensions by the MPC in concert with an athlete. We interpreted that survey use is not necessarily contrary to narrative approaches. The idea that surveys can facilitate dialogue is not novel; however, the notion that many MPCs wished to embed items and scales within a conversation, as part of a think-aloud to ascertain metacognitive/motivational information from a client, is different than a more sequenced ‘assess first’, then ‘analyze and discuss second’ approach. This approach blends dialogue and survey items in exchanges between an MPC and a client with the goal of ‘assessment as learning’ rather than ‘assessment of learning’. Overall, we suggest survey use could be beneficial and innovative, perhaps for many more consultants, were a less stringent psychometric approach ascribed to assessment. Such an approach implicates MPCs using surveys in personalized, integral, and reasoned ways. Notably, several of our seasoned MPCs expressed that they believed they could work through the SRSP content by relying on their prerogatives and own expertise to extend into dialogue. This is an important observation because expert MPCs like to rely on their accumulated experience in their own work to guide aspects of service delivery [38]. Based on many of our MPCs’ responses, the SRSP appears to provide a foundation which is not constraining, from which experienced MPCs believe they can adequately enlarge dialogue based on their personal expertise.

When quantitative surveys are construed by MPCs in monolithic terms (e.g., exclusively psychometric diagnosis), the prospective uses of survey content may be significantly diminished or dismissed. Moreover, survey use cannot be based on anecdotal intuitions in the absence of evidence-based methods. Our results are important because they offer a preliminary, prospective evidence base for different ways of using surveys, apart from preconceived/stereotyped notions related to testing and clinical diagnosis. This evidence permits MPCs to think of client-centered ways to integrate a survey into consulting, strategically, to prompt qualitative needs analysis. The evidence invites a person-centric approach in which survey use is personalized by the MPC and tailored to the client’s needs. These findings may therefore be insightful for MPCs who have been trained in recent decades to develop more client-led, flexible, values-oriented consulting accorded to humanistic philosophies [e.g., 38,39,40], for whom a client’s needs analysis is predicated upon “a range of methods” [41, p. 506), featuring “a qualitative type of description” [p. 507], but commonly omitting surveys.

For those in our camp of Canadian MPCs who preferred quantitative survey use, the scores mattered. They mattered for potentially providing metrics that could support tracking within an unfolding trajectory of consulting. Scores also mattered for prompting dialogue during needs analyses and gathering information for case conceptualization. With respect to this latter facet, the prospective goal of these MPCs was not to use surveys to simply obtain scores/metrics, rather to ensure rich dialogue based on scores/metrics. Thus, our evidence shows the use of scores within applied consulting is very different than positivist conceptions of scores, which are concerned with minimizing threats to the objective reality of data [e.g., 42].

### Initial Findings Contributing to the Prospective Practical Validity of The SRSP Survey

Sport psychology scholars have lamented a research-to-practice gap [43], reflecting the disconnect between what evidenced-based empirical work provides and how practitioners can actually use such evidence in real-world practice [44]. This disconnect is attributed to many factors, including the inability to reconcile the needs/observations of practitioners with the theoretical/measurement concerns of academics, the lack of input from practitioners into academic researchers’ designs, a predominant emphasis in academic research on internal validity, failure of researchers to produce works that are intended for the practitioner, and the under-reporting or over-complicating of implementation recommendations [43,44]. Winter et al. [41] interviewed British sport psychologists about the processes of evidence-informed decision making around methods in applied sport psychology. The practitioners expressed that the sport psychology literature commonly lacks the types of research that are most useful in aiding their applied decision-making processes and therefore, they are not inclined to consider methods arising from this work. Because published quantitative surveys typically originate in an academic setting with goals for testing, practitioners have concerns that these instruments neglect how the tools may be used in real-world practice where goals are different.

The SRSP was designed from academic work, for empirical purposes, that did not originate with practitioners [19]. Much of the early research leading to the SRSP survey [e.g., 24,26,27] was concerned with measurement/psychometrics and/or testing whether self-report scores could reliably discriminate more-elite groups of athletes from less-elite groups. This academic research was successful in determining an elite-advantage around SRSP scores, leading to the conclusion that better training habits and enhanced development of elite athletes could be attributed to their strategic engagement in constituent SRSP proficiencies [18]. However, it remains uncertain whether the SRSP survey could be of utility to MPCs in supporting the development of individual athletes, and specifically in a dyadic consulting context.

The findings show initial evidence supporting the prospective use of the SRSP among professional MPCs. Our results show that Canadian MPCs who have an interest in working on psychological proficiencies to enhance practice are open to using the SRSP survey and see potential value in its use. The exception is consultants who have philosophical reservations of using any type of quantitative survey. Furthermore, we found that most of the Canadian MPCs believed that the survey could benefit them in early phases of case conceptualization with a client, which involves assessment and analysis, interpretation of strengths and weaknesses, and enlargement to understand an athlete’s needs around training, and various scenarios and contextual features. The MPCs’ responses suggested that use of the SRSP could vary from traditional quantitative assessment with psychometric underpinnings to more innovative, exploratory narrative inquiry, all personalized to an MPC’s consulting philosophy. Overall, the findings are an initial step that establishes the prospective practical validity of the SRSP survey.

The current findings are an extension of the perspectives of Canadian MPCs offered by Young et al. [33]. In that article, researchers noted that consultants were aware of the topic of SRL and could identity with it in their practice (i.e., the ‘what’ of psychology of practice). The MPCs also described the merits of adolescent athletes being self-regulated learners in terms of quality practice (the ‘why’ of psychology of practice). That article tendered self-regulated sport practice as a worthwhile and attractive area of consultancy that MPCs were interested in targeting with adolescents - a form of practical validity. The current findings add to this evidence by locating the SRSP survey as a prospective instrumental asset for some MPCs.

Our findings align with a case study in which a Polish MPC used a SRSP survey to engage a client in a reflective exercise on the development of practice habits [16]. By asking an international-level athlete to complete the survey while reflecting on their habits from five years ago, then redoing the same exercise asking them to reflect on their habits today, the MPC opened up a narrative on growth whereby she could reinforce metacognitive strengths for the athlete. To our knowledge, this is the lone study that animates a specific exercise leveraging the SRSP survey in consulting. The current findings further contribute to dialogue supporting the SRSP among practitioners. One of our key research questions was ‘what information do MPCs believe they can gain by giving the SRSP to clients?’. We interpret a two-fold answer. First, the MPCs who were inclined to use the SRSP said they could gain information benefitting their role in case conceptualization. Secondly, they could describe ways in which they might begin to act on information drawn from survey-related discussion as they move towards analyzing and developing an athlete’s proficiencies. On the latter point, however, more detail on specific actionable steps, prescription of psychological skills, and structured aspects of intervention that follow from SRSP-narratives is needed to elaborate on practical utility. Moreover, moving beyond prospective to authentic practical validity requires research involving actual use of an athlete’s SRSP responses, exploration of whether these responses reflect the athlete’s own understandings, and whether and how an MPC can interpret, analyze and act on responses effectively with that athlete. It is possible that the mock exercise may have encouraged participants to imagine idealized responses rather than the difficulties that arise when working with real clients; as a result, the current accounts may be more optimistic than practitioners’ actual behavior when employing the survey in an authentic applied setting. Still, we believe that the current findings are a first step in starting a conversation to ‘narrow the gap’ between the theoretical/measurement works on the SRSP and its implementation in the field.

### Discerning Autonomy Support, Perceived Autonomy and Self-Regulated Sport Practice Proficiencies

Another of our key questions inquired about MPCs’ perceptions of the psychological content being assessed by the SRSP. In their descriptions, they recognized metacognitive, motivational and SRL content, which is faithful with the conceptual background of the survey [19]. More broadly, the MPCs’ interpretations of content suggest that the SRSP could be useful for discussion of triadic interactions between: environmental constraints on self-directedness; an athlete’s qualities of ownership, feelings of independence and self-leadership; and an athlete’s capability to proficiently self-manage aspects of their training.

Autonomy is a ubiquitous term within sport psychology literature. Certainly, autonomy is an integral psychological need for adolescent athletes supported by vast research within self-determination theory [see 45]. However, the sport psychology literature sometimes espouses autonomy with little discernment for the environment, the person, and the psychological properties that the athlete needs to ‘step into’ autonomy. Our finding that many MPCs wish to use the SRSP to figure out whether a client has opportunities to self-direct within their training environment is connected to the notion that different sport cultures (e.g., gymnastics versus canoe/kayak) and different coaching styles (a very coach-directed/coach-controlled style versus an “autonomy-supportive” style) [46] may constrain/open *opportunities* for self-regulated sport practice learning and application. For example, Radowits and Young [47] found variability in how adolescent sport coaches afforded “time and space” for prompting and reinforcing adolescent athletes’ experimentation with SRL around training. These affordances are important and intersect with an athlete’s *perceptions of autonomy* in various times or spaces where they might exercise self-regulated sport practice. The finding that some of our MPCs would use the SRSP to figure out the extent to which an athlete felt a sense of ownership and cognitive engagement in their own learning can be inserted in this facet. Finally, even when an environment affords opportunities to exercise autonomy, and even when an athlete perceives autonomy, it is not guaranteed that they will act skillfully. Thus, we submit that self-regulated sport practice proficiencies may be the requisite psychological properties that equip young athletes to act skillfully in an autonomy-supported environment when they feel they can exercise autonomy. These properties are necessary because affordances and perceptions are not sufficient to optimize skill acquisition for sport talent development. Young people are expected to vary in how skillfully they apply such properties and, as clients, they may benefit by gaining these proficiencies as outcomes of future applied interventions. The current study is limited in that it neither operationalizes the nature of these proficiencies nor the specific actionable strategies that support their development. These should be aims of future work in SRSP-related research.

### Limitations and Future Research

In line with pragmatism, we present this article as an overture to inquiry on the psychology of practice, involving relevant practitioners, and tender the findings as provisional knowledge that should be subject to future revision by other researchers [30]. Findings are limited to a Canadian context, to a self-selected sample of MPCs that was restricted to practitioners already interested in consulting on practice enhancement. This was appropriate for eliciting informed views, but it also means that we cannot provide an account of practitioner receptivity across the wider profession. These MPCs participated in Young et al. [33], which meant they had considered the concept of SRL in an earlier part of their lengthy interview before being asked about the SRSP, which could have resulted in more coherent responses. Whereas we prompted the consideration of clients via a mock SRSP, future research should include authentic, in-person interactions with clients. Research is also needed to examine what real athlete clients think about the survey and what they believe they need from MPCs in SRSP-related protocol.

The current study looked at initial and global perspectives on the SRSP survey. Future studies are required to understand how MPCs plan to support and engage a client in meaning-making for each of the five constituent scales of the SRSP, how these meanings relate to experiences/scenarios in sport training, and how the MPCs would specifically intervene to support psychological skill development. The current article presented findings limited to early phases of case conceptualization and client analysis (hence the importance of enlargement) and therefore did not fully consider other aspects of dyadic consulting. For instance, case conceptualization ultimately directs subsequent selection stages of sport psychology consulting, including prescriptions/actions supporting psychological skill training [1]. Moreover, case conceptualization is an iterative process whereby the profile of a client and initial working hypotheses for intervention are constantly updated as an intervention unfolds [1,48]. How the SRSP survey and SRSP-related narratives interact with such considerations will be important to address in future research. Finally, some of our MPCs mentioned surveys they already use in response to issues around practice, such as the OMSAT-3 [20], TOPS-2 [21], and PCDEQ [49]. It would be beneficial to explore what Wilson et al.’s [19] SRSP survey offers uniquely and complementarily, to practitioners, relative to these established resources.

## Author Information

* Bradley W. Young, School of Human Kinetics, University of Ottawa, Ottawa, ON Canada, K1N 6N5; Lisa Bain, School of Human Kinetics, University of Ottawa, Ottawa, ON Canada, K1N 6N5; Sharleen Hoar, Coastal Human Performance, Victoria, BC Canada V8N 5V6; Joseph Baker, Tanenbaum Institute for Science in Sport, University of Toronto, ON Canada, M5S 2W6. 0000-0002-5686-1737.

## Declarations

The authors declare that there are no personal, financial, or institutional conflicts of interest regarding the research in this article. The writing of this manuscript was funded by an Insight Grant from the Social Sciences and Humanities Research Council of Canada (435-2018-1481).

## Data Availability

No quantitative data were generated by this study. All data collection was qualitative and transcribed verbatim. The research ethics certificate affiliated with this study does not allow for public/open access to participants' transcripts. We have disclosed all relevant excerpts in the manuscript. We are able to share additional excerpts in support of interpretations upon request, however, we do not have informed consent to share transcripts in a pubic repository.

